# Prevalence of Tobacco Pouch Keratosis in India: A Systematic Review and Meta-Analysis Protocol

**DOI:** 10.1101/2025.08.27.25334605

**Authors:** Chandru Sivamani, Kavipriya Outtamane, Sombuddha Biswas, S Kavya, Mamta Mor, Revathy Ajayan, Sooriyaprasannan Sriram

## Abstract

**Background:** Tobacco pouch keratosis (TPK) is a common oral mucosal lesion among smokeless tobacco users, with potential for malignant transformation. Despite its public health importance, pooled prevalence estimates for India remains limited.

**Methods:** A comprehensive search will be conducted in PubMed, Embase, and Scopus, along with grey literature sources (Google Scholar and Shodhganga). Observational epidemiological studies published from 2000 onwards, reporting the prevalence of TPK in India, will be included. Forward and backward citation searches will also be performed. The study selection process will be documented using the PRISMA 2020 flow diagram. Data will be extracted using the Joanna Briggs Institute (JBI) standardized form, and methodological quality will be assessed using the JBI Critical Appraisal Checklist for prevalence studies. Key study characteristics and results will be tabulated, and provide a narrative synthesis. Pooled prevalence will be estimated using a random-effects model (restricted maximum likelihood method), with heterogeneity assessed via Cochran’s Q and I^2^ statistics. If fewer than five studies are available, the Hartung– Knapp–Sidik–Jonkman method will be applied. Subgroup analyses will be performed by age group, gender, and study setting. Publication bias will be evaluated using funnel plots and Egger’s test. The certainty of evidence will be graded using the GRADEpro framework.

**Discussion:** This review will generate pooled prevalence estimates of TPK across diverse populations and settings in India and will also provide evidence-based insights for researchers, clinicians, public health professionals, and policymakers involved in tobacco control and oral health initiatives.

**PROSPERO registration:** CRD420251119683

## Introduction

Smokeless tobacco (SLT) use has become a major public health concern over the recent decades, with more than 360 million users worldwide, the majority residing in South Asia.[1] In 2020, about 27% of all diagnosed cancer cases in India were attributable to tobacco use.[2] According to the World Health Organization (WHO), by 2030 tobacco consumption is projected to cause over 8 million deaths annually, accounting for 10% of global mortality.[3] Furthermore, tobacco-related deaths exceed the combined mortality of the three major communicable diseases (CDs) such as tuberculosis (TB), human immunodeficiency virus (HIV) or acquired immunodeficiency syndrome (AIDS), and malaria. Tobacco Pouch Keratosis (TPK), a localized oral potentially malignant disorder (OPMD) characterized by hyperkeratotic mucosal changes due to habitual placement of SLT in the buccal vestibule, which can progressively develop into a thickened keratotic plaque with distinct pouch formation, potentially progressing to oral squamous cell carcinoma.[4] Despite the implementation of tobacco control measures, the national level surveys (Global Adult Tobacco Survey (GATS) and the Global Youth Tobacco Survey (GYTS) have reported a high prevalence of SLT use among individuals aged ≥13 years.[5,6] This raises the significant concern about the growing burden of TPK and other OPMDs. Although TPK is clinically distinct, it carries a risk of malignant transformation, and its epidemiological burden remains underexplored, particularly in the Indian context. While numerous studies have reported the prevalence of tobacco-associated oral mucosal lesions in different populations, data specific to TPK are fragmented and geographically inconsistent.[7,8] To date, no comprehensive systematic review and meta-analysis has synthesized the prevalence estimates for TPK in India, despite the country’s high SLT burden. Therefore, this review aims to synthesize the available evidence on TPK prevalence in India, providing a robust pooled estimate to support disease surveillance, promote early diagnosis, guide targeted health interventions, and reinforce tobacco control policies.

## Methods

The systematic review and meta-analysis (SRMA) on the prevalence of tobacco pouch keratosis (TPK) in India was prospectively registered in PROSPERO on May 8, 2025 (CRD420251119683), and any future amendments to the protocol will be updated in the PROSPERO record. The protocol has been developed in accordance with the Preferred Reporting Items for Systematic Review and Meta-Analysis Protocols (PRISMA-P) 2015 statement (**Supplementary file 1**).[9] This SRMA will be conducted in accordance with the relevant chapter of the JBI manual for evidence synthesis.[10]

### Eligibility criteria

The review will include studies that report the prevalence of TPK among the Indian population, published from January 1, 2000, to ensure the inclusion of epidemiologically relevant data that reflects current patterns of smokeless tobacco use, diagnostic practices, and reporting standards. Recent studies reported high prevalence estimates of TPK, ranging from 15% to 46.1%.[4,7,8,11] Studies conducted in the early 2000s may not adequately capture these patterns due to changes in tobacco product formulations, public health awareness, and healthcare-seeking behavior. The CoCoPop (Condition, Context, and Population) framework guided the inclusion criteria for this review.[10] To be eligible for inclusion, studies must address the relevant condition (tobacco pouch keratosis), provide data within the appropriate context (prevalence), and focus on the target population (Indian population).

### Condition (Tobacco Pouch Keratosis)

Tobacco pouch keratosis is characterized by a white, corrugated plaque on the oral mucosa at the site where smokeless tobacco is habitually placed. Early lesions may present as a thin white film, progressing over time to a thicker keratotic lesion with distinct pouching. Histologically, these lesions often display hyperkeratotic and/or acanthotic squamous epithelium, sometimes with epithelial dysplasia.[4,12] For this review, cases will be included if diagnosed clinically by a dental professional, with or without histopathological confirmation, as reported in the original studies.

### Context (Prevalence)

We will include the observational epidemiological studies such as cross-sectional, longitudinal studies (baseline data), and case-control studies (only healthy group) that report the prevalence data on TPK. However, we will exclude the intervention studies, systematic reviews, case series, case reports, and editorials from this review.

### Target population (Indian population)

The target population for this review will include the individuals residing in India from various geographic locations (urban, rural, and tribal areas), both males and females aged ≥13 years (GATS-3 and GYTS-4),[5,6] and any occupational groups who use smokeless tobacco products and have been diagnosed with TPK will be included.

### Search strategy

Searches will be conducted in multiple databases, including PubMed, Embase, and Scopus, along with gray literature sources such as Google Scholar and Shodhganga. The initial search will be performed on October 1, 2025, and updated in January 2026 prior to the final analysis. Search strategies will be developed using the CoCoPop framework, incorporating relevant MeSH terms and keywords related to TPK, disease measurements, and the Indian population, adapted to the functionality of each database (**Supplementary file 2**). Boolean operators (AND, OR) will be used in combination with controlled vocabulary and free-text terms, as appropriate. Forward and backward citation tracking of relevant reviews will also be undertaken to identify additional eligible studies.

### Data management

The search results from all databases will be exported in RIS/nbib format and imported into Rayyan (https://www.rayyan.ai/), a web-based tool for screening and managing systematic review records.[13] Rayyan will automatically identify duplicate records, and we will manually remove the duplicates before the screening process. Two reviewers will independently screen titles and abstracts in Rayyan, blinded to each other’s decisions. Each record will be marked as “include,” “exclude,” or “maybe,” and exclusion reasons will be recorded for excluded studies. After the initial screening, decisions will be unblinded, and conflicts will be resolved through discussion, with a third reviewer acting as arbitrator if required. Studies marked as “include” or “maybe” will proceed to full-text screening within Rayyan, where the same process of independent, blinded assessment and conflict resolution will be followed. The study selection process will be documented and presented in accordance with the Preferred Reporting Items for Systematic Reviews and Meta-Analyses (PRISMA) 2020 flow diagram. Final included studies will then be exported from Rayyan for data extraction (Microsoft Excel) and analysis (RStudio version 4.4.2 or STATA). The Grading of Recommendations, Assessment, Development and Evaluation (GRADE) approach will be applied to assess the certainty of evidence, and the summary of findings tables will be reported using the GRADEpro web application (https://www.gradepro.org/).[14]

### Data extraction

Data will be extracted from all studies that meet the eligibility criteria after a full-text review. To ensure consistency, we will use a standardized data extraction form, adapted from the JBI manual (**Supplementary file 3**).[10] The extracted variables will include: (i) bibliographic and descriptive information (such as authors, title, year of publication, and study location); (ii) methodological parameters (including study design, study setting, study population, inclusion and exclusion criteria, sample size, diagnostic definitions (clinically or histopathological, and type of smokeless tobacco); and (iii) outcome data (prevalence with 95% confidence intervals, and/or standard errors)

### Risk of bias assessment

The methodological quality and risk of bias in the included studies will be evaluated using the JBI critical appraisal checklist for studies reporting prevalence data.[10] The tool consists of nine items that evaluate potential sources of bias, including: (1) appropriateness of the sample frame to address the target population; (2) appropriateness of the sampling method; (3) adequacy of sample size; (4) detailed description of study subjects and setting; (5) coverage of the identified sample in the analysis; (6) validity of methods used to identify the condition; (7) reliability and standardization of measurement; (8) appropriateness of statistical analysis; and (9) adequacy and management of the response rate.

Each item will be rated as “Yes,” “No,” “Unclear,” or “Not applicable.” For synthesis, “Yes” responses will be scored as 1 and all other responses as 0. The total score will be calculated by summing the number of “Yes” responses and dividing by the total number of applicable items. Studies will be classified as having low risk of bias (≥75% “Yes” responses; ≥7/9), moderate risk (50–74%; 4–6/9), or high risk (<50%; ≤3/9). Two reviewers will conduct the appraisal independently, and disagreements will be resolved by discussion or consultation with a third reviewer. To facilitate visual interpretation, the risk-of-bias assessments will also be presented using a traffic light system, where “Yes” will be represented by the colour green (indicating low risk for a domain), “Unclear” will be represented by the colour yellow (indicating insufficient information), and “No” will be represented by the colour red (indicating high risk for that domain). An overall traffic light summary table will be prepared, displaying each study’s ratings across all nine JBI items, along with the final risk-of-bias classification.[10,15]

### Data synthesis

Based on the inclusion and exclusion criteria, a table summarizing the study characteristics and results will be prepared to highlight the key details for each study, and also a narrative synthesis will be presented to summarize the relevant studies, supported by a meta-analysis to compute the pooled estimates of the prevalence of TPK in both community-based and hospital-based settings. We will employ the random effects maximum likelihood method for the meta-analyses, given the expected heterogeneity from study samples across different national populations in India.[16,17] If the number of included studies is less than five, we will use the Hartung-Knapp-Sidik-Jonkman method to estimate the pooled prevalence of TPK.[18]

Subgroup analyses will also be performed with Cochran’s Q and Higgins’ I^2^ used to test for heterogeneity among different age groups(13–24, 25–44, and ≥45 years) in line with the classification used in major national surveys: the Global Youth Tobacco Survey (GYTS) for youth (13–15 years), the Global Adult Tobacco Survey (GATS) for adults (≥15 years, often split into 15–24 and 25–44 age groups), and the National Family Health Survey (NFHS) for ages 15–49.[5,6,19] The subgroup analyses will also consider factors such as gender, location, and study design (cross-sectional, retrospective, and longitudinal).

### Meta-bias(es)

Statistical heterogeneity will be evaluated using Cochran’s Q test and Higgins’ I^2^ statistic. In cases where ten or more studies are available, potential publication bias will be examined through visual inspection of funnel plots and tested using Egger’s regression method.[20] Sensitivity analyses will be performed to assess the robustness of the pooled prevalence estimates.

### Confidence in cumulative evidence

The overall certainty of the evidence will be assessed and summarized using the GRADE approach. Although there is limited specific guidance on applying GRADE to prevalence-focused systematic reviews, it remains the recommended framework, with some relevant guidance available in the literature.[21,22]

## Discussion

Tobacco pouch keratosis remains an under-recognized oral lesion in India, despite its well-documented association with smokeless tobacco use and potential for malignant transformation.[23,24] Existing literature on TPK prevalence is fragmented, with variations in the study settings and populations, making it difficult to obtain a reliable national estimate. This systematic review and meta-analysis will address these gaps by synthesizing epidemiological data from diverse settings and subpopulations, thereby providing more precise prevalence estimates. The planned subgroup analyses by age group, gender, and study setting will allow a better understanding of demographic and contextual factors influencing TPK distribution. Employing rigorous screening, critical appraisal using the JBI tool, and quantitative synthesis with robust statistical methods will enhance the reliability of the findings. The use of the GRADE framework will further ensure transparent evaluation of evidence certainty. Potential limitations include heterogeneity in study methodologies, possible underreporting in certain populations, and publication bias. To overcome these, comprehensive database and grey literature searches will be undertaken, along with sensitivity analyses and bias assessments. The results from this review are expected to inform oral health surveillance, guide early detection and prevention strategies, and support tobacco control policy formulation in India. By pooling the available evidence, the review will provide a valuable resource for researchers, clinicians, and public health authorities aiming to reduce the burden of TPK and related oral cancers.

## Data Availability

All data produced in the present study are available upon reasonable request to the authors

## Author contributions

**Conceptualization:** Chandru Sivamani, Kavipriya Outtamane, Sombuddha Biswas, Kavya S, Mamta Mor, Revathy Ajayan, Sooriyaprasannan Sriram; **Methodology:** Chandru Sivamani; **Writing – Original Draft:** Chandru Sivamani; **Writing – Review & Editing:** Chandru Sivamani, Kavipriya Outtamane, Sombuddha Biswas, Kavya S, Mamta Mor, Revathy Ajayan, Sooriyaprasannan Sriram; All authors approved the final version of the manuscript, and agree to be accountable for all aspects of the work. Chandru Sivamani acts as the **guarantor** for this work.

## Availability of data and materials

Not applicable

## Declarations

### Ethical approval and consent to participate

Not applicable

