## Supplementray file 2 for "Prevalence of Tobacco Pouch Keratosis in India: A Systematic Review and Meta-Analysis Protocol"

Search strategy for all the data base

PubMed search strategy:

| #1 | (((((((((((((((((((((tobacco pouch keratosis[Title/Abstract]) OR (tobacco induced keratosis[Title/Abstract])) OR (tobacco use disorder[Title/Abstract])) OR ("keratosis"[MeSH Terms])) OR (Kerato*[Title/Abstract])) OR (white keratotic lesion[Title/Abstract])) OR (smokeless tobacco keratosis[Title/Abstract])) OR (smokeless tobacco associated keratosis[Title/Abstract])) OR (oral mucosal lesion*[Title/Abstract])) OR (oral premalignant condition*[Title/Abstract])) OR (oral premaligna*[Title/Abstract])) OR (oral premalignant lesion*[Title/Abstract])) OR (snuff dippers lesion[Title/Abstract])) OR (oral potentially malignant lesions[Title/Abstract])) OR (oral potentially malignant disorder*[Title/Abstract])) OR (smokeless tobacco types[Title/Abstract])) OR ("tobacco, smokeless"[MeSH Terms])) OR (tobacco consumption[Title/Abstract])) OR (tobacco induced oral changes[Title/Abstract])) OR (tobacco chewing habit*[Title/Abstract])) OR (tobacco related lesion[Title/Abstract])) OR (tobacco related lesion*[Title/Abstract]) |
| --- | --- |
| #2 | ((((((((((prevalence[Title/Abstract]) OR (prevalence[MeSH Terms])) OR (frequency[Title/Abstract])) OR (distribution[Title/Abstract])) OR (incidence[Title/Abstract])) OR (incidence[MeSH Terms])) OR (epidemiology[Title/Abstract])) OR ("epidemiology"[MeSH Terms])) OR (epidemiologic studies[Title/Abstract])) OR (occur*[Title/Abstract])) OR ("epidemiologic studies"[MeSH Terms]) |
| #3 | (("india"[Title/Abstract]) OR (india*[Title/Abstract])) OR (india[MeSH Terms]) |
| #4 | #1 AND #2 AND #3 |

Embase search strategy:

| 1# | ('tobacco pouch keratosis':ti,ab OR 'tobacco induced keratosis':ti,ab OR 'tobacco use disorder':ti,ab OR 'keratosis':ti,ab OR kerato*:ti,ab OR 'white keratotic lesion':ti,ab OR 'smokeless tobacco keratosis':ti,ab OR 'smokeless tobacco associated keratosis':ti,ab OR 'oral mucosal lesion*':ti,ab OR 'oral premalignant condition*':ti,ab OR 'oral premaligna*':ti,ab OR 'oral premalignant lesion*':ti,ab OR 'snuff dippers lesion':ti,ab OR 'oral potentially malignant lesions':ti,ab OR 'oral potentially malignant disorder*':ti,ab OR 'smokeless tobacco types':ti,ab OR 'tobacco smokeless':ti,ab OR 'tobacco consumption':ti,ab OR 'tobacco induced oral changes':ti,ab OR 'tobacco chewing habit*':ti,ab OR 'tobacco related lesion':ti,ab OR 'tobacco related lesion*':ti,ab) |
| --- | --- |
| #2 | (prevalence:ti,ab OR frequency:ti,ab OR distribution:ti,ab OR incidence:ti,ab OR epidemiology:ti,ab OR 'epidemiologic studies':ti,ab OR occur*:ti,ab) |
| #3 | ('india':ti,ab OR india*:ti,ab) |
| #4 | #1 AND #2 AND #3 |

Scopus search strategy:

| 1# | TITLE-ABS-KEY ("tobacco pouch keratosis") OR ("tobacco induced keratosis") OR ("tobacco use disorder") OR (keratosis) OR (kerato*) OR ("white keratotic lesion") OR ("smokeless tobacco keratosis") OR ("smokeless tobacco associated keratosis") OR ("oral mucosal lesion*") OR ("oral premalignant condition*") OR ("oral premaligna*") OR ("oral premalignant lesion*") OR ("snuff dippers lesion") OR ("oral potentially malignant lesions") OR ("oral potentially malignant disorder*") OR ("smokeless tobacco types") OR ("tobacco smokeless") OR ("tobacco consumption") OR ("tobacco induced oral changes") OR ("tobacco chewing habit*") OR ("tobacco related lesion") OR ("tobacco related lesion*") |
| --- | --- |
| #2 | TITLE-ABS-KEY (prevalence) OR (frequency) OR (distribution) OR (incidence) OR (epidemiology) OR ("epidemiologic studies") OR (occur*) |
| #3 | TITLE-ABS-KEY (india) OR (india*) |
| #4 | #1 AND #2 AND #3 |

Google Scholar search strategy:

| #1 | ("tobacco pouch keratosis" OR "oral potentially malignant lesions" OR "oral potentially malignant disorder*" OR "tobacco induced oral)  AND (prevalence OR frequency OR distribution OR incidence OR epidemiology OR "epidemiologic studies" OR occur*) AND (india) |
| --- | --- |
