## Supplementray file 3 for "Prevalence of Tobacco Pouch Keratosis in India: A Systematic Review and Meta-Analysis Protocol"

| Sl.no | Author | Year | State | Journal name | Article title | Funding | Conflict | Study design | Study setting | Time period | Study population | Inclusion criteria | Exclusion criteria | Sample size | Age (mean ±SD) | Male  (n%) | Female  (n%) | Socio  economic  status | Prevalence  (95% CI) | Diagnostic criteria |
| --- | --- | --- | --- | --- | --- | --- | --- | --- | --- | --- | --- | --- | --- | --- | --- | --- | --- | --- | --- | --- |
| Study 1 |  |  |  |  |  |  |  |  |  |  |  |  |  |  |  |  |  |  |  |  |
| Study 2 |  |  |  |  |  |  |  |  |  |  |  |  |  |  |  |  |  |  |  |  |
| Study 3 |  |  |  |  |  |  |  |  |  |  |  |  |  |  |  |  |  |  |  |  |
| Study 4 |  |  |  |  |  |  |  |  |  |  |  |  |  |  |  |  |  |  |  |  |
| Study 5 |  |  |  |  |  |  |  |  |  |  |  |  |  |  |  |  |  |  |  |  |

Supplementary file 3: Data extraction from
